# Malaria prevalence and its associated factors amongst rural adults: Cross-sectional study in East Nusa Tenggara Province Indonesia

**DOI:** 10.1101/2024.09.11.24313521

**Authors:** Robertus Dole Guntur, Jusrry Rosalina Pahnael, Keristina Br Ginting, Yulianti Paula Bria, Damai Kusumaningrum, Fakir M Amirul Islam

**Affiliations:** Mathematics Study Program, Faculty of Science and Engineering, Nusa Cendana University, Kupang NTT, Indonesia; Department of Computer Science, Widya Mandira Catholic University Kupang NTT, Indonesia; Animal Health Study Program, Department of Animal Husbandry State Agricultural Polytechnic, Kupang NTT, Indonesia; Department of Health Science and Biostatistics, School of Health Sciences, Swinburne University of Technology, Melbourne, Australia

**Keywords:** Malaria prevalence, risk factors, rural adults

## Abstract

**Introduction:** Malaria is a global health issue including in Indonesia. Currently, most of the cases was in the rural of eastern part of the country. However, malaria risk factors amongst rural adult were less documented. This study investigated malaria risk factors amongst rural adults in East Nusa Tenggara Province (ENTP).

**Methods:** A community based cross-sectional study was conducted to interview 1495 rural adults in ENTP. A multi-stage cluster random sampling technique was applied to collect data on malaria history, demographic, behavioural and environmental factors of malaria of participants. Logistic regression model was applied to decide significant factors associated with malaria.

**Results:** The prevalence of malaria was 13.4%. The prevalence of malaria was significantly higher for adults with low level of malaria knowledge (Adjusted odd ratio (AOR): 2.43, 95% confidence interval (CI): 1.38 – 4.27) compared to those with high malaria knowledge level, having moderate malaria knowledge level (AOR: 1.99, 95% CI: 1.11 – 3.57) compared to those high malaria knowledge level, having no education (AOR: 2.18, 95% CI: 1.37 – 3.45) compared to those junior high school or above education level, outdoor occupation (AOR: 1.81, 95% CI: 1.22 – 2.68) compared to indoor occupation, having family size > 4 (AOR 2.08, 95% CI: 1.52 –2.87) compared to those ≤ 4

**Conclusion:** The study revealed the prevalence of malaria amongst rural adults in this province was high. The study highlights the power of malaria knowledge level on the prediction of malaria prevalence amongst rural adults. Health education intervention is critical for vulnerable groups to reduce malaria prevalence in the province.

## Introduction

Malaria is a major global health problem, with an estimation that in 2022, there were 3.4 billion people living at risk of contracting malaria, and approximately 1.23 billion of them are at high risk to have malaria [1]. Based on current World Health Organization (WHO) data, the number of malaria cases in 2022 was around 2.8 million; 2 % of this amount came from Southeast Asia (SEA) countries. The action plan in the Southeast Asia region shows that by 2030 the entire region would be expected to be malaria free [2]. Of the 11 countries in SEA, WHO has awarded a certificate as malaria-free country to the Maldives and Sri Lanka. One of the indicators for obtaining this certificate is that there must be no local malaria cases in the last three consecutive years [2]. India and Indonesia still suffer from malaria, accounting for 65.7% and 22.4% of the total number of malaria cases in the region in 2022 [1].

Following the global commitment, Indonesia is under progress to become a malaria-free region before 2030 [3,4] Much progress has been made to support this international commitment. From the total of 514 districts in the country, 75.7% percent of these have been classified as malaria-free districts in 2023 [5]. However, there is a large gap between the Western and Eastern Parts of Indonesia in achieving this. The majority of districts in the western part of this country have been classified as malaria-free areas, while only a few districts in the eastern region of the Republic of Indonesia, including in the Province of East Nusa Tenggara (ENT) have achieved malaria-free district status [5].

ENT Province, which is the seventh largest contributor to all national malaria cases in 2023, has 22 districts [5]. Forty-one percent of the districts in this archipelagic province have been classified as malaria-free areas, and other districts are categorized as high, moderate and low malaria endemic areas. The number of malaria cases has shown a downward trend in the last decade [6], but during the Covid-19 pandemic, the trend of malaria cases increased significantly, from 12,909 in 2019 to 16,219 in 2022 [7]. Even worse, the number of deaths rose significantly during that period from 2 deaths in the first year of Covid-19 in 2020 to 8 deaths at the end of Covid-19 period in 2022 [8]. The distribution of this fatality rate is uniform in four districts of Kupang, South Central Timor, East Sumba and Southwest Sumba, two cases in each district. The parasite aspect of malaria in this province reveals that plasmodium falciparum was the common causes of malaria in this province, however, the prevalence of plasmodium vivax is much high in this region [6,9,10]. The existence of plasmodium vivax provide unique challenge for the effort of malaria elimination as the effectiveness of treatment require high awareness of community for consuming the medicine for 14 days [11,12].

Malaria is a complex problem and is associated with many factors including plasmodium types [13–15], environmental factors [16–18]; climate change [19–21]; behavioural aspects [22]; sociodemographic factors [23–25]. Moreover, the occurrence of malaria might shows significantly different trend within and across countries because of variation in micro-epidemiological factors such as genetic, social, environmental, and other factors [26]. The current systematic review of investigating malaria risk factors in Southeast Asia by exploring database study from the period of 2010 to 2020 in this region indicated that the malaria risk factors study is commonly only focus on environmental and weather aspects of malaria [27]. To boost malaria elimination, behaviour aspect including local knowledge of malaria should be incorporated in modelling of the malaria transmission to understand well the distribution trend of the disease in community level [28].

Many studies have been conducted in Indonesia to investigate the potential factors associated with malaria prevalence in this country. A study of the spatial distribution of malaria prevalence carried out in Papua Province discovered that rural Papuan staying in poor and lowland districts were the high risk to have malaria [29]. Analysis on active and passive surveillance data from Aceh Besar, shows that the likelihood to have malaria for adults having 16 – 45 years of age was 14 times higher than those ≤15 years [30]. Other studies indicated that prevalence of plasmodium vivax and falciparum was higher in female than their counterpart and being a farmer [31], raising medium-size animal in the house [32], and low level of education were significantly associated with malaria [24]. However, those studies did not consider socio economic, demographic, behavioural and environmental aspects of malaria simultaneously. Better understanding of malaria transmission might be achieved by exploring malaria risk factors comprehensively [17].

Furthermore, in the ENT Province, the investigation on malaria risk factors on the behavioural aspect at village level indicated that behaviour of local community was associated with incidence of malaria in the Kori Vilage [33]. Analysis of environmental aspect of malaria in this area indicated that the decline of malaria prevalence of all age group due to long distance from the larva habitat [34] and there is a strong association between malaria infection and surrounding of the local community [35]. Socio-demographic data analysis of urban and rural community in this province reveals that the risk of malaria of low education level was higher than their counterpart [36]. However, those studies apply small samples and covering urban-rural population. The research on malaria human aspects for rural community was less documented in this area, despite the significant contribution of malaria knowledge of human to progress to malaria elimination [37,38] and the high burden of malaria was contributed by rural population [4]. Human behaviour and community perspective is imperative for malaria elimination [39] and their level on malaria knowledge should be measured and considered in implementing various program to reduced malaria prevalence [40]. The current literature indicates that malaria awareness and malaria knowledge of rural community in ENT Province was very low [41,42]. However, the effect of malaria knowledge on the malaria infection has not been investigated yet in the study area. Developing a malaria prevalence model by considering many factors including the malaria knowledge level is very important to be able to provide the best model in determining the best predictors of malaria infection. Therefore, the present study aimed to fill this gap by exploring the effect of malaria knowledge level and other factors in developing malaria risk factors of rural communities of ENT Province to support the local and national government in achieving malaria elimination by 2030.

## Materials and Methods

### Study sites and period of study

The Province of ENT is an archipelago province located in the eastern part of Indonesia. The province consists of 21 districts and one municipality spreading cross six main island including Flores, Sumba, Timor, Alor, Lembata and Sabu Raijua island [43]. The province has a total population about 5.6 million with the almost balance proportion between male and female. Most of population work in agricultural sector. This community based cross-sectional study was conducted from September to October 2019 in three district including East Sumba, East Manggarai and Belu district. The study area has two main seasons, namely dry season from June to September and rainy season from December to March. The average temperature in this area is 27 – 28^0^C with the lowest at 15.8^0^C and the highest at 32.8^0^C [43].

### Sample size calculation

The initial sample size (*n*_0_) was computed by applying the formula for prevalence study in a cross-sectional study [44] with formula :

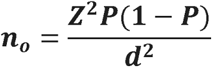

Where parameter Z indicates the value of standard score of 95% confidence (1.96), P describes the prevalence of malaria research in ENT Province previously done by Health Ministry of Indonesian government (1.99%) [45], and d reveals the relative precision namely 0.01125. Therefore, the initial sample size equals to 592. By considering design effect because of cluster sampling interval, the initial sample size (n_o_) was multiplied by a factor 2.16. The final sample size (n) was computed by taking into account 85% of participation rate of the study resulting, *n* = 1503 participants. The completed calculation of sample size was shown in the previous study [46].

### Sampling technique and data collection method

A multi-stage cluster sampling method with a systematic random sampling technique at the final cluster level 4 was employed for recruiting rural adults from three different districts. Firstly, at the cluster level one, three districts were chosen based on their malaria endemic settings (MES), from high, low and moderate MES. Secondly, at the cluster level two, three sub-districts were randomly nominated from each selected district. Thirdly, at the cluster level three, a number of villages was selected by considering their relative population. Finally, at the cluster level four, a systematic random sampling method was applied to enrol 20 – 40 participants in each village. One adult over 18 years of age of any gender was selected from each selected household. As we want to estimate the prevalence of malaria for rural adults, we exclude anyone who is under 18 years of age. In each visited household, a participant was interviewed guided by a structure questionnaire conducted by local nurses who had participated in the intensive training of data collection prior this study. The process of data collection was scrutinized by the principal investigator of the study on daily basis for monitoring the completeness and the accuracy of the data in the questionnaire.

### Outcome Variables

The dependent variable of the study was self-reported malaria and it is a kind of binary variable which is whether malaria is absent or present. This variable was derived from a question whether he/she had been diagnosed malaria in the past 24 months by local health care providers having tools to confirmed malaria in their facilities. A dichotomous variable coded as one if participant answer yes to this question and zero if she/he reported otherwise.

There is no screening test during the survey as we applied “self-reported malaria” as outcome variable. However, in coordination with local health centres, all respondents reported malaria history in the last two year were checked their malaria history in the malaria register book of the centre.

### Socio-demographic covariates

The independent variables of the study were grouped into three categories including socio- demographic, environmental dan behavioural aspects of malaria. Socio-demographic variables were gender classified as male and female, age group categorized into five group which is < 30, 30 – 39, 40 – 49, 50 – 59, and > 60 years of age. Education level of participants was classified as no education, primary education level, and junior high school level or above. Family size was grouped into four or less than four family member and greater than four family members. The main occupation of participants has two categories which is indoor and outdoor occupation. Social economic status (SES) of participants was classified as low, average, and high SES. Furthermore, the environmental aspects of malaria in this study consist of the distance to nearest health facilities classified as < 1 km, 1 - 2 Km, > 2 Km; modern house of participants was classified as yes and no; source of drinking water was categorized as running tap water dan open source, and the access to TV was grouped into yes and no. Moreover, the behavioural aspects of malaria comprise as the use of bed net (yes or no), poor treatment seeking behaviour, and the level of malaria knowledge. Poor treatment seeking behaviour was defined as the previous publication [12]. Participants having perception to seek malaria treatment within 24 hours and at health facilities were categorized as good malaria treatment seeking behaviour, whilst for those seeking treatment after 24 hours or seeking treatment at non health facilities were classified to have poor appropriate malaria treatment seeking behaviour for malaria. The level of malaria knowledge of rural community was classified as high, moderate and low level as indicated in the previous study [41].

### Statistical Analysis

The initial stage in conducting data analysis for cross-sectional data was carried out using the descriptive analysis method [47,48]. This method was used to see the distribution of participants based on demographic, environmental and behavioral aspects of malaria. Then we apply chi-square test to see the variation of malaria prevalence amongst variable predictors and to investigate the initial association amongst variable predictors and dependent variable. Data analysis was continued by modeling using a logistic regression model. All variables showing a significant association in the chi-square test were included in the bivariate logistic regression. The use of traditional levels, such as P value at 0.05 might fail to select variables known to be imperative, we applied a P cut-off point of 0.10 in the Wald test in the bivariate analysis [49].

In the multivariable analysis, after controlling the confounding variables, all variables with a p-value of 0.05 or less were considered statistically significant as a predictor of malaria prevalence. The direction and strength of association between explanatory variables and endpoints were estimated by adjusting the odds ratio. Statistical software SPSS version 27 (SPSS Inc.) was used for analyses.

### Ethics approval

The research was performed in accordance with the tenets of The Declaration of Helsinki. The study was accepted by the Human Ethics Committee of Swinburne University of Technology, Australia (Reference: 20191428–1490) and the Health Research Ethics Committee, National Institute of Health Research and Development (HERC-NIHRD), Ministry of Health of Indonesia (Reference: LB.02.01/2/KE.418/2019).

## Results

The participation rate of the study was high (99.5%). There were eight participants missing information, therefore the data of 1945 participants were included in this analysis. The characteristics of participants based on the socio-demographic, environmental and behavioural factors were presented in Table 1. Socio-demographic characteristic of 1495 participants revealed that the percentage of female participants was 51.4%, with 95% confidence interval (CI): 47.8- 54.9, almost half of participants (45.4% with 95% CI: 41.7 - 49.1) completed their primary school, more than half of participants (62.9% with 95% CI: 59.8, 66.0) was outdoor occupation, and more than half of them (57.5%, with 95% CI: 54.2, 60.8) had average socio-economic status. With regard to behavioural factors, the percentage of participant having low level of malaria knowledge was 51.2% with 95% CI: 47.7, 54.7, and it was 48% with 95% CI: 44.4, 51.7 with poor understanding of appropriate malaria treatment seeking behaviour, whilst the percentage of participants using bed net in the night prior to the survey was high, which is 80.7% with 95% CI: 78.5, 82.9. In relation to environmental factors, only 24% (95% CI: 19.8, 28.6) of participants lived in modern house, 59.3% (95% CI: 56.0, 62.5) of them used opensource water, and 38.7% (95% CI: 34.7, 42.7) of participants resided less than one kilometres from the nearest health facilities.

**Table 1.** Distribution of participants amongst rural adults in East Nusa Tenggara Province, Indonesia (N=1495)

| Characteristics | n | % | 95% CI |
| --- | --- | --- | --- |
| Overall | 1495 |  |  |
| **Gender** |  |  |  |
| Male | 727 | 48.6 | [45.0, 52.3] |
| Female | 768 | 51.4 | [47.8, 54.9] |
| **Age group** |  |  |  |
| < 30 | 205 | 13.7 | [9.00, 18.4] |
| 30 -39 | 418 | 28.0 | [23.7, 32.3] |
| 40 - 49 | 371 | 24.8 | [20.4, 29.2] |
| 50 - 59 | 295 | 19.7 | [15.2, 24.3] |
| > 60 | 206 | 13.8 | [9.07, 18.5] |
| **Level of education** |  |  |  |
| No Education | 279 | 18.7 | [14.1, 23.3] |
| Primary School | 678 | 45.4 | [41.7, 49.1] |
| Junior High School or above | 538 | 36.0 | [31.9, 40.1] |
| **Main Occupation** |  |  |  |
| Indoor | 554 | 37.1 | [33.1, 41.1] |
| Outdoor | 941 | 62.9 | [59.8, 66.0] |
| **Socio-Economic Status** |  |  |  |
| Low | 449 | 30.0 | [25.8, 34.3] |
| Average | 860 | 57.5 | [54.2, 60.8] |
| High | 186 | 12.4 | [7.70, 17.2] |
| **Family size** |  |  |  |
| < = 4 | 803 | 53.7 | [50.3, 57.2] |
| > 4 | 692 | 46.3 | [42.6, 50.0] |
| **†HH income in relation to PMW** |  |  |  |
| < PMW | 1342 | 89.8 | [88.1, 91.4] |
| >= PMW | 153 | 10.2 | [5.43, 15.0] |
| **Distance to the nearest health service** |  |  |  |
| < 1 Km | 578 | 38.7 | [34.7, 42.7] |
| 1 - 2 Km | 400 | 26.8 | [22.5, 31.1] |
| > 2 Km | 517 | 34.6 | [30.5, 38.7] |
| **Modern House** |  |  |  |
| No | 1133 | 75.8 | [73.3, 78.3] |
| Yes | 362 | 24.2 | [19.8, 28.6] |
| **Source of drinking water** |  |  |  |
| Running tap water | 609 | 40.7 | [36.8, 44.6] |
| Open source | 886 | 59.3 | [56.0, 62.5] |
| **Access to TV** |  |  |  |
| Yes | 582 | 38.9 | [35.0, 42.9] |
| No | 913 | 61.1 | [57.9, 64.2] |
| **Malaria knowledge level** |  |  |  |
| High | 260 | 17.4 | [12.8, 22.0] |
| Moderate | 470 | 31.4 | [27.2, 35.6] |
| Low | 765 | 51.2 | [47.7, 54.7] |
| **Poor appropriate treatment seeking behaviour** |  |  |  |
| No | 718 | 48.0 | [44.4, 51.7] |
| Yes | 777 | 52.0 | [48.5, 55.5] |
| **The use of bed net** |  |  |  |
| Yes | 1207 | 80.7 | [78.5, 82.9] |
| No | 288 | 19.3 | [14.7, 23.9] |
| **Having travel history in the last one month** |  |  |  |
| Yes | 83 | 5.6 | [0.65, 10.5] |
| No | 1412 | 94.4 | [93.2, 95.6] |

### The variation of malaria prevalence amongst independent variables

The variation of malaria prevalence amongst various factors including demographic, environmental, and behavioural aspects of malaria was presented in Table 2. Overall, the prevalence of malaria was 13.4%. The malaria prevalence was not statistically difference amongst participants with different gender (p = 0.34) and age group (p = 0.508). However, a statistically significant disparity was noticed in malaria prevalence amongst participant having different education level (p < 0.001). Malaria prevalence was the highest in the group of participants with no education (25%), whilst it was the lowest in the group with junior high school or above (9.9%). Furthermore, the variation of malaria prevalence was significant statistically amongst participant with different main occupation (p = 0.001), where the proportion of participants having malaria prevalence was the higher in the group of outdoor occupation (15.7%) than in the group of indoor occupation (9.57%). Moreover, the results showed that there was a significant discrepancy in malaria prevalence amongst participants with diverse malaria knowledge level (p <0.001). The highest prevalence of malaria was observed in the group of participants with low level of malaria knowledge (16.3%), whilst it was the lowest in the group of people with high level of malaria knowledge (6.15%).

**Table 2.**
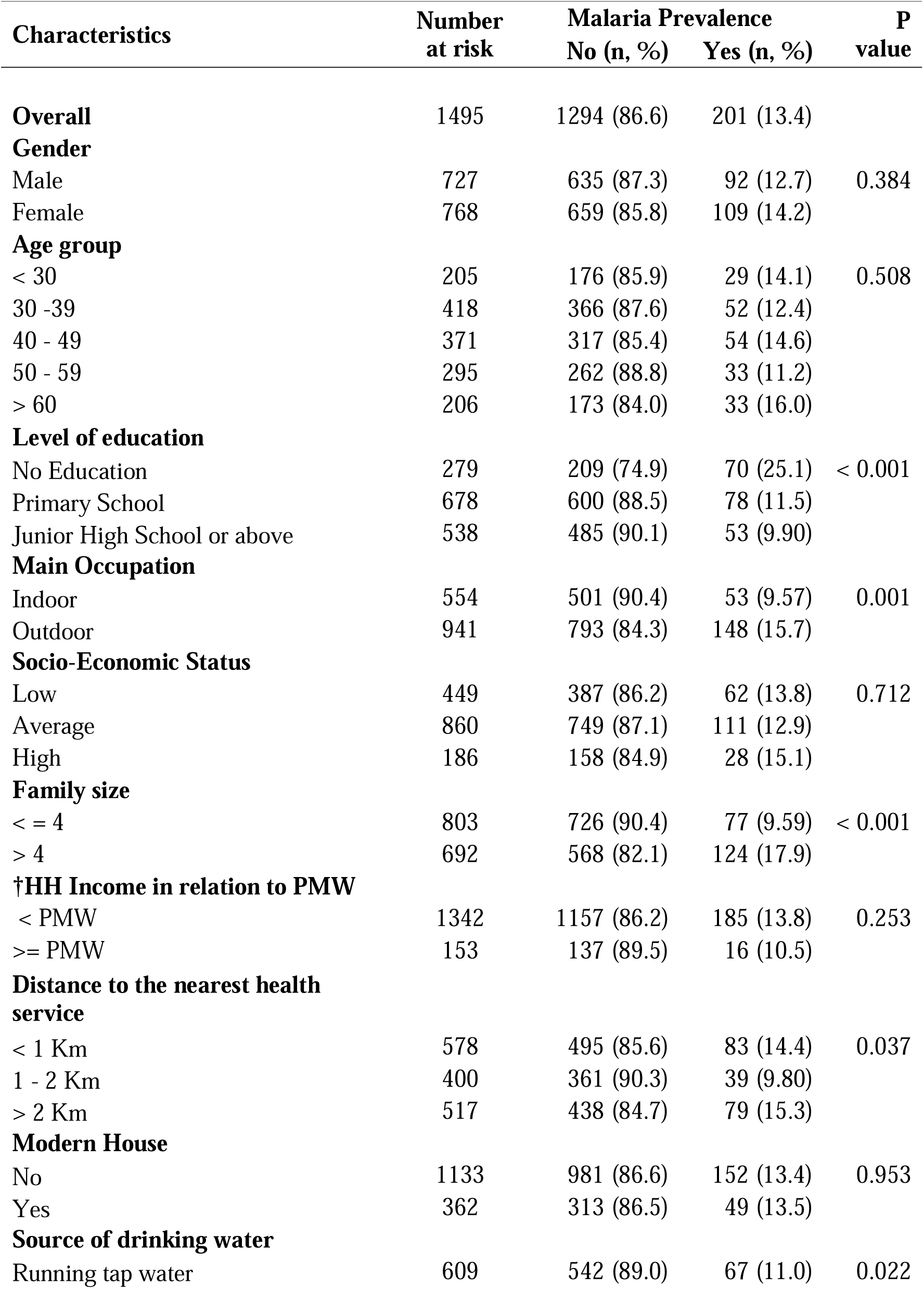
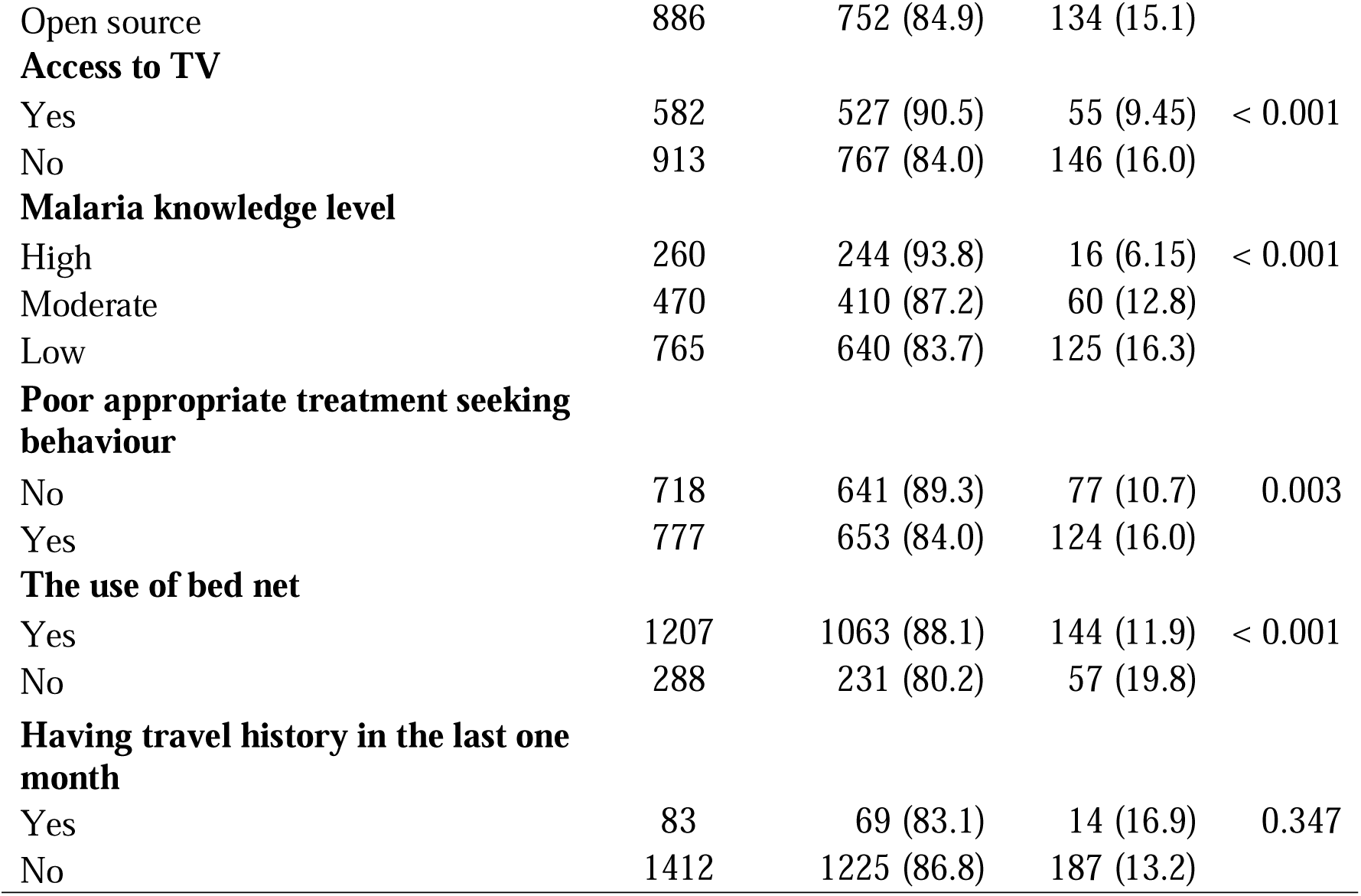
The association of demographic, environmental and behavioural factors with malaria prevalence amongst rural adults in East Nusa Tenggara Province, Indonesia (N=1495)

The association between malaria prevalence and variable independents in bivariate analysis was presented in Table 3. The bivariate logistic regression demonstrated that participants having no education level were more likely to report malaria relative to those having junior high school level or above (Odd ratio (OR): 3.07, 95% confidence interval (CI): 2.07 – 4.54). By main occupation, it was evidence that the likelihood of malaria prevalence among rural adults with outdoor occupation was almost two times higher than those with indoor occupation (OR: 1.76, 95% CI :1.26 – 2.46). Rural adults having family size greater than four were two times higher to have malaria prevalence than those having family size less than four (OR: 2.06, 95% CI: 1.52 – 2.79). In terms of environmental factors, it was clear that source of drinking water and access to TV shows a significant association with malaria prevalence. Rural adults having open-source drinking water were more likely to report malaria relative to those having running tap water (OR: 1.44, 95% CI: 1.05 – 1.97), then the likelihood of malaria prevalence among rural adults having no access to TV was almost two times higher than those having access to TV (OR: 1.82, 95% CI: 1.31 – 2.54). In terms of behavioural aspect of malaria, it was clear that malaria knowledge level, poor appropriate treatment seeking behaviour, and the use of bed net had a significant association with malaria prevalence. Rural adults having low level of malaria knowledge were almost three times higher to have malaria than those high level of malaria knowledge (OR: 2.98, 95% CI: 1.73 – 5.12). Participants having poor appropriate treatment seeking behaviour were more likely to report malaria relative to those their counterpart (OR: 1.58, 95% CI: 1.17 – 2.14). Rural adults who did not use the bed net were almost two times higher to have malaria than those using bed net (OR: 1.82, 95% CI: 1.30 – 2.55) as indicated in Table 3.

**Table 3.** Multivariable binary logistic regression: factors associated with malaria prevalence amongst rural adults in East Nusa Tenggara Province, Indonesia.

| **Characteristics** | **Malaria prevalence** |  |
| --- | --- | --- |
|  | **COR (95% CI) \*** | **AOR (95% CI) \*\*** |
| **Gender** |  |  |
| Male | 1.00 (Reference) | 1.00 (Reference) |
| Female | 1.14 (0.85, 1.54) | 1.32 (0.93, 1.88) |
| **Age group** |  |  |
| < 30 | 1.00 (Reference) | 1.00 (Reference) |
| 30 -39 | 0.86 (0.53, 1.41) | 0.70 (0.41, 1.17) |
| 40 - 49 | 1.03 (0.64, 1.68) | 0.80 (0.47, 1.35) |
| 50 - 59 | 0.76 (0.45, 1.30) | 0.60 (0.34, 1.07) |
| > 60 | 1.16 (0.67, 1.99) | 0.77 (0.42, 1.40) |
| **Level of education** |  |  |
| No Education | 3.07 (2.07, 4.54) | 2.18 (1.37, 3.45) |
| Primary School | 1.19 (0.82, 1.72) | 0.94 (0.63, 1.42) |
| Junior High School or above | 1.00 (Reference) | 1.00 (Reference) |
| **Main Occupation** |  |  |
| Indoor | 1.00 (Reference) | 1.00 (Reference) |
| Outdoor | 1.76 (1.26, 2.46) | 1.81 (1.22, 2.68) |
| **Family size** |  |  |
| < = 4 | 1.00 (Reference) | 1.00 (Reference) |
| > 4 | 2.06 (1.52, 2.79) | 2.08 (1.52, 2.87) |
| **Distance to the nearest health service** |  |  |
| < 1 Km | 1.00 (Reference) | 1.00 (Reference) |
| 1 - 2 Km | 0.64 (0.43, 0.97) | 0.68 (0.45, 1.03) |
| > 2 Km | 1.08 (0.77, 1.50) | 1.03 (0.73, 1.46) |
| **Source of drinking water** |  |  |
| Running tap water | 1.00 (Reference) | 1.00 (Reference) |
| Open source | 1.44 (1.05, 1.97) | 1.37 (0.99, 1.89) |
| **Access to TV** |  |  |
| Yes | 1.00 (Reference) | 1.00 (Reference) |
| No | 1.82 (1.31, 2.54) | 1.36 (0.96, 1.94) |
| **Malaria knowledge level** |  |  |
| High | 1.00 (Reference) | 1.00 (Reference) |
| Moderate | 2.23 (1.26, 3.96) | 1.99 (1.11, 3.57) |
| Low | 2.98 (1.73, 5.12) | 2.43 (1.38, 4.27) |
| **Poor appropriate treatment seeking behaviour** |  |  |
| No | 1.00 (Reference) | 1.00 (Reference) |
| Yes | 1.58 (1.17, 2.14) | 1.19 (0.85, 1.65) |
| **The use of bed net** |  |  |
| Yes | 1.00 (Reference) | 1.00 (Reference) |
| No | 1.82 (1.30, 2.55) | 1.39 (0.97, 1.98) |

\*COR: Crude odds ratio, \*\*AOR: Adjusted odds ratio, CI: confidence interval

### Multivariable Analysis

The adjusted association between malaria prevalence and other aspects including socio- demographic, environmental and behavioural aspects of malaria was depicted in Table 3. After adjustment for all covariates variables, the following factors were associated with malaria prevalence: low and moderate malaria knowledge, no education level, outdoor occupation and large family size. Rural adults having low malaria knowledge level were 2.43 times higher to have malaria prevalence than those having high malaria knowledge level (Adjusted Odd Ratio (AOR): 2.43, 95% CI: 1.38 – 4.27) and rural adults with moderate malaria knowledge level were almost two times higher to have malaria prevalence than those having high malaria knowledge level (AOR: 1.99, 95% CI: 1.11– 3.57). By education level, it was evident that rural adults with no education level were two times higher to have malaria than those with junior education level or above (AOR: 2.18, 95% CI: 1.37 – 3.45). Moreover, rural adults having family size greater than four were two times higher than those with less than four (AOR: 2.08, 95% CI: 1.52 – 2.87). The likelihood of malaria prevalence among rural adults with outdoor occupation was almost two times higher than those with indoor occupation (AOR: 1.81, 95% CI: 1.22 – 2.68). Even though in the bivariate analysis variables including access to TV, the use of bed net, poor appropriate treatment seeking behaviour, and source of drinking water show a significant association, in the final model those variables were not significant statistically as shown in Table 3.

## Discussion

This study addresses a research gap in malaria prevalence and its associated risk factors amongst rural adults in ENTP Indonesia. The major finding of the study was the malaria prevalence was prevalent in the study area. The main factors associated with the prevalence of malaria were low and moderate level of malaria knowledge, low level of education, outdoor occupation, big family size. These implies that to progress to malaria elimination in this region, health education intervention is critical to improve knowledge of local community about malaria prevention measures and control strategy and find out additional vector control strategies targeting outdoor malaria transmission in the rural community of this province. Whilst, other factors including socio-economic status, distance to the nearest health facilities, access to television, source of drinking water, and the use of bed net did not demonstrate a significant association with malaria prevalence in this study.

This study demonstrates that the level of malaria knowledge was a significant factor associated with malaria prevalence. Malaria prevalence amongst rural adults with low level of malaria knowledge was two times higher than those with high level of malaria knowledge and in the group of moderate malaria knowledge level, their malaria prevalence was almost two times higher than their counterparts. This finding consistent with other studies indicating that low level of malaria knowledge rose the possibility to have malaria infection [50–53]. This could be explained by the fact that people with low level of malaria knowledge have no idea on how to recognize the symptom on the disease, how to prevent the disease, when and where to seek the appropriate treatment during their sick. Furthermore, other studies indicated that the variation of malaria prevention measure knowledge was low in this study area [54] and the knowledge on long-lasting insecticide-treated nets (LLINs), which is the most effective prevention measures to prevent malaria[38,55], was only common on the high endemic settings in this province [56]. Moreover, another study revealed that the practice of using non-LLINs was only common in low endemic setting [57]. These situations could provide more challenge on the effort to reduce malaria prevalence in this area. Improving malaria knowledge of local community in low, moderate and high endemic settings in this province could increase chance to practice of various malaria prevention measures, leads to reduce malaria prevalence [58] and to boost malaria elimination [59].

The present study verified that there was a significant association between education level and malaria prevalence. The prevalence of malaria amongst rural adults decreased significantly with the increase of education level of participants. It was noticed that rural adult with no education level had the highest prevalence rate, whilst the lowest was in the group of adults with junior high school or above education level. This finding was consistent with other studies in Indonesia [60–62] and other studies globally [63–66] demonstrating the power of high education level to reduce the prevalence of malaria. This could be explained by the fact that people with high level of education had a skill to understand written concept of information [67] and had many chance to be exposure with multiple source of information [68]. As a result, this group of people have could identify the symptoms of the disease, how to prevent the diseases, where to seek appropriate treatment [12] which in turn could reduce the prevalence of malaria [64]. This emphasized the role of education to boost malaria elimination in the study area. The local authority should gear the policy to improve the education level of the rural community to reduce the burden of malaria in this region.

The finding of this research also indicated that outdoor occupation was significantly associated with malaria prevalence. This study shows that malaria prevalence of rural adults having outdoor occupation was almost two times higher compared to those having indoor occupation. This result was consistent with finding of other studies [50,69,70] indicating the highest prevalence of malaria amongst those with outdoor job. This might be the fact that people with outdoor activities are exposed with outdoor biting of *Anopheles* mosquitoes which is quite prevalent in the study area [71]. Moreover, the majority of rural community in this study area are engaged in agricultural sectors [42] enabling them to have high chance to be bitten by the mosquitoes. Improving awareness of this group of people to protect themselves during their work is critical to boost malaria elimination in this region.

This study further showed that there is a considerable association between family size and malaria prevalence. The study has shown that malaria prevalence for rural adults having family size more than four was two times higher compared to those having family size less than four. Our finding corroborates with studies in other settings [72–74] indicating the positive correlation between the malaria prevalence and the number of people in a house. The more people in house, the higher the risk of contracting malaria. This could be understood as crowding’s potent smell signals draw in more mosquitoes [75].

This study highlights the level of malaria knowledge as a main predictor for malaria prevalence in this region. Therefore, it is suggested that to accelerate for the achievement of malaria elimination certificate in this province, the partnership amongst sectors is critical. This includes collaboration between education and health department in developing malaria intervention program to enhance malaria knowledge of local rural community. Education department should encourage school to have malaria knowledge class in their local curriculum. The great achievement of China government to gain malaria elimination zone in 2021 because of massive effort to improve malaria awareness of student’s community [76,77]. Moreover, literature indicated that the prevalence of lower education [78] and drop out of school children [79,80] was high in ENT Province. Intervention for improving malaria knowledge level of rural community by speaker announcement [81] and music [82] might work well in this area as indicated in other countries.

The current study employed a big sample size incorporating rural adults aged from 18 years to 89 years from the general population of Indonesia’s ENTP, enabling a comprehensive understanding of malaria prevalence in the rural communities in the country. However, there are some weaknesses of the research, for example, malaria prevalence was computed on the history of malaria reported by participants. Moreover, due to the nature of cross-sectional study, malaria prevalence was only computed only in the specific period, therefore the trend and the pattern of the malaria prevalence could not be depicted by study. Further investigation could possibly be done by combining data from medical record of patient in the local and national level of the country to present the change pattern of malaria prevalence from time to time. Despite these drawbacks, this study provides insight about malaria situation and risk factors in the effort of the country to achieve of malaria elimination by 2030.

## Conclusions

The study revealed the prevalence of malaria amongst rural adults in this province was high. Low and moderate level of malaria knowledge, no education, outdoor occupation and big family size were the important risk factors of malaria. The study emphasizes the power of malaria knowledge level on the prediction of malaria prevalence amongst rural adults. Therefore, health education intervention to improve the level of malaria knowledge is critical for those groups to reduce malaria prevalence in the province.

## Data Availability

All data produced in the present study are available upon reasonable request to the authors

## Acknowledgments

We would like to express our gratitude to the Health Ministry of Indonesia, the governor of ENTP, head of East Sumba, Belu, and East Manggarai district, nine head of sub-districts, and forty-nine village leaders for allowing this research to be performed in their regions.

